# Insights on improving accessibility and usability of functional data to unlock its potential for variant interpretation

**DOI:** 10.1101/2025.01.25.25321117

**Authors:** Min Seon Park, Runjun D. Kumar, Cristian Ovadiuc, Andrew Folta, Abbye E. McEwen, Ashley Snyder, Douglas M. Fowler, Alan F. Rubin, Brian H. Shirts, Lea M. Starita, Andrew B. Stergachis

## Abstract

**Introduction:** Variant-level functional data is a core component of clinical variant classification and can aid in reinterpreting variants of uncertain significance (VUS). However, the usage of functional data by genetics professionals is currently unknown.

**Methods:** An online survey was developed and distributed in spring of 2024 to individuals actively engaged in variant interpretation. Quantitative and qualitative methods were used to assess responses.

**Results:** 190 eligible individuals responded, with 93% reporting interpreting 26 or more variants per year. The median respondent reported 11-20 years of experience. The most common professional roles were laboratory medical geneticists (23%) and variant review scientists (23%). 77% reported using functional data for variant interpretation in a clinical setting and overall respondents felt confident in assessing functional data. However, 67% indicated that functional data for variants of interest was rarely or never available, and 91% considered insufficient quality metrics or confidence in the accuracy of data as barriers to its use. 94% of respondents noted that better access to primary functional data and standardized interpretation of functional data would improve usage. Respondents also indicated that handling conflicting functional data is a common challenge in variant interpretation that is not performed in a systematic manner across institutions.

**Discussion:** The results from this survey showed a demand for a comprehensive database with reliable quality metrics to support use of functional evidence in clinical variant interpretation. The results also highlight a need for guidelines regarding how putatively conflicting functional data should be used for variant classification.

## Introduction

Precise genetic diagnosis is a pillar of genomic medicine^1,2^. However, many variants identified by genetic testing lack the evidence needed to be classified as pathogenic or benign, rendering them variants of uncertain significance (VUS)^3,4^. This lack of evidence predominantly impacts missense variants, with over 90% of the 1.1 million unique variants in ClinVar currently classified as VUS^5^. Approximately 50% of clinically reported variants are VUS^6,7^ and they have a profound impact on genetics providers and their patients because they cannot be used in diagnosis or guiding treatment^8–11^. Moreover, the burden of VUS is not equally shouldered by different populations^12,13^. Consequently, each test that returns with a VUS is a missed opportunity to provide optimal genomic medicine.

Evidence for resolving a VUS classification to pathogenic/likely-pathogenic (P/LP) or benign/likely-benign (B/LB) can come from patient phenotype, segregation analysis, population frequency, *in silico* variant effect predictions, or experimental functional data^4^. Experimental assays can range from simple *in vitro* enzymatic assays to mouse models of specific human variants. More recently, multiplexed assays for variant effect (MAVE) have been deployed to generate functional data at scale^14^. In a MAVE, nearly all possible variants of a target sequence can be tested in a functional assay in parallel, enabling the functional data generated by a MAVE to serve as a look-up table for variant function in perpetuity^14–16^. The Clinical Genome Resource Sequence Variant Interpretation Working Group (ClinGen SVI) has established guidelines for translating functional data into PS3/BS3 strong/moderate/supporting evidence for variant classification^17^. And when available, the addition of functional data to other evidence has been shown to enable reclassification of 15-75% of VUS for commonly tested tumor suppressor genes like *BRCA1, TP53, PTEN* and *MSH2*^18,19^. Furthermore, because all possible variants can be included in the functional assay, MAVEs have been shown to reduce the bias in VUS rate between individuals of different ancestries^20^. However, functional data can be hard to find, difficult to interpret, and are the single largest contributor to conflicting variant interpretations between clinical laboratories^21^.

To better understand the needs of the international genomic medicine community in advancing broad use of functional data, we sought to: (1) explore how providers were encountering and reinterpreting VUS; (2) assess provider experience and comfort using functional data and related resources; and (3) identify barriers that providers perceive are preventing the use of functional data. We also sought direct feedback on best practices for communicating functional data and metadata alongside other standard variant-specific information. We developed and deployed a needs assessment survey to gather this information, soliciting responses across the global genetics community. Responses from a diverse set of genome medicine professionals reveal that VUS are encountered often, and most are not reinterpreted. Many attempt to use functional data as evidence for VUS but are unsure how to apply the data or resolve conflicting information. Respondents identified attributes of a structured resource to disseminate functional data that would help overcome these barriers.

## Materials & Methods

### Survey design

To inform the survey design, we conducted informal interviews with two genetic counselors, a molecular geneticist at a private diagnostic company, a board-certified laboratory geneticist at an academic medical center, a clinical molecular geneticist in the US government, and a board-certified medical geneticist at a private healthcare company. These interviews were used to refine the scope and language used in the survey questions and identify the intended recipients.

The study was designed as a REDCap survey (**Supplemental Information**), which consisted of 79 questions (Likert-scale, multiple choice, and free-text response questions) that included sections addressing: (1) variant interpretation and reinterpretation practice, (2) current and anticipated professional role, (3) comfort level with various functional evidence categories, guidelines, and resources, (4) dealing with conflicting functional evidence data, (5) barriers and facilitators to using functional evidence data in variant interpretation, (6) attitudes toward proposed functional data dashboard, and (7) demographic information. It was anticipated that this survey would take 10-15 minutes to complete. The sole inclusion criteria was that respondents should be actively engaged in interpreting genetic variants for clinical significance (whether or not these interpretations are ultimately reported to patients). We anticipated that this would include clinical professionals such as clinical geneticists, clinical laboratory geneticists, genetic counselors, and variant review scientists, as well as some researchers.

### Survey distribution

The survey was distributed by e-mail or newsletter to members of the following organizations whose members would be anticipated to meet our criteria: (1) American Board of Medical Genetics and Genomics (ABMGG); (2) National Society of Genetic Counselors (NSGC); (3) Association for Molecular Pathology (AMP); (4) the Clinical Genetics Resource (ClinGen); and (5) CanVIG-UK. Additionally, the survey was directly advertised during seminars and in between sessions at international conferences such as the 2024 American College of Medical Genetics annual meeting, the 2024 European Society for Human Genetics annual meeting, the 2024 Mutational Scanning Symposium, and the 2024 Association for Clinical Genomic Science annual meeting. Across email and newsletter recipients and conference attendees, we estimate more than 10,000 people could have been invited to respond to the survey, however membership in each group is highly overlapping and an exact number is impossible to calculate. The first survey response was collected on February 8th of 2024, and the last survey response was collected on June 18th of 2024, with 50% of the survey responses being collected between February 8th and March 6th of 2024.

### Survey analysis

199 individuals submitted a response to this survey, 9 were excluded due to not meeting eligibility criteria of being involved with clinical variant interpretation. One participant did not provide an answer to the question asking if they are involved in variant interpretation for clinical significance. However, this respondent noted that they interpret 6-25 variants per year, indicating that they are involved with variant interpretation. Consequently, that individual was deemed to meet the eligibility criteria, and their responses were included in the analysis. Data from all of these 190 respondents were used for the analyses. Aside from the question asking about the respondents’ involvement with clinical variant interpretation to determine eligibility, none of the questions were required to complete the survey. All of the Likert-scale and Yes/No questions had at least 182/190 (96%) responses. 139/190 (73%) of respondents completed all of the Likert-scale survey questions, with 184/190 (97%) completing at least 90% of the Likert-scale questions. Data was collected and stored through REDCap, and analyzed using R (RStudio Version 2024.09.1+394). For sub-analyses that involved ranking responses from individual respondents, if a respondent reported the same comfort level for all categories, their response was excluded from this specific sub-analysis. Figures were generated using R (RStudio Version 2024.09.1+394) and refined with Adobe Illustrator (version 29.2. 1). R code utilized for analyses and raw data are available at https://doi.org/10.5281/zenodo.14735953^22^.

Some demographic data has been removed to avoid potential participant identification. Qualitative data from free text responses were uploaded to Dedoose software (version 9.2.22), and the responses for handling conflicting functional evidence with 116 responses were analyzed using inductive thematic analysis^23,24^. Other free-response sections with less than 20 responses were analyzed for patterns. A member of the research team (MSP) coded the responses and identified emerging themes and patterns. Some responses were assigned more than one of the identified themes, reflecting multifaceted content of the response. The identified themes were presented and reviewed by the research team.

## Results

### Survey captures responses from a diverse set of clinical genetics professionals

We distributed an online survey (**Supplementary Information**) to clinical and research genetics professionals, with the goal of better understanding how genomic medicine providers interacted with VUS and functional data. In all, a total of 190 individuals who responded to the survey met the eligibility criteria. We achieved international representation with a majority of respondents indicating the United States as their primary country of practice (63%), followed by the United Kingdom (17%), and 15 different countries across five continents (**Figure 1A**). Approximately half of the respondents were early career professionals with less than 10 years of experience (48%, **Figure 1B**) but all career stages were well represented. Respondents work in a variety of settings, with academic medical centers being the most common (46.3%, **Figure 1C**). The three most common professional roles were laboratory medical geneticist (23.1%), variant review scientist (22.6%), and clinical MD geneticist (17.9%) (**Figure 1D**). More than half of the respondents perform variant interpretation for pathogenicity in clinical diagnostic settings (79%) as well as return of genetic testing results to patients (63%, **Figure 2A**). In addition, the majority responded that they are currently involved with curating functional evidence for clinical variant classification (67%) and using functional evidence to reassess variant classification in a clinical setting (77%, **Figure 2B**). Overall, this indicates that this survey captured a diverse set of medical professionals who are actively involved in clinical variant interpretation and actively use functional data.

**Figure 1.**
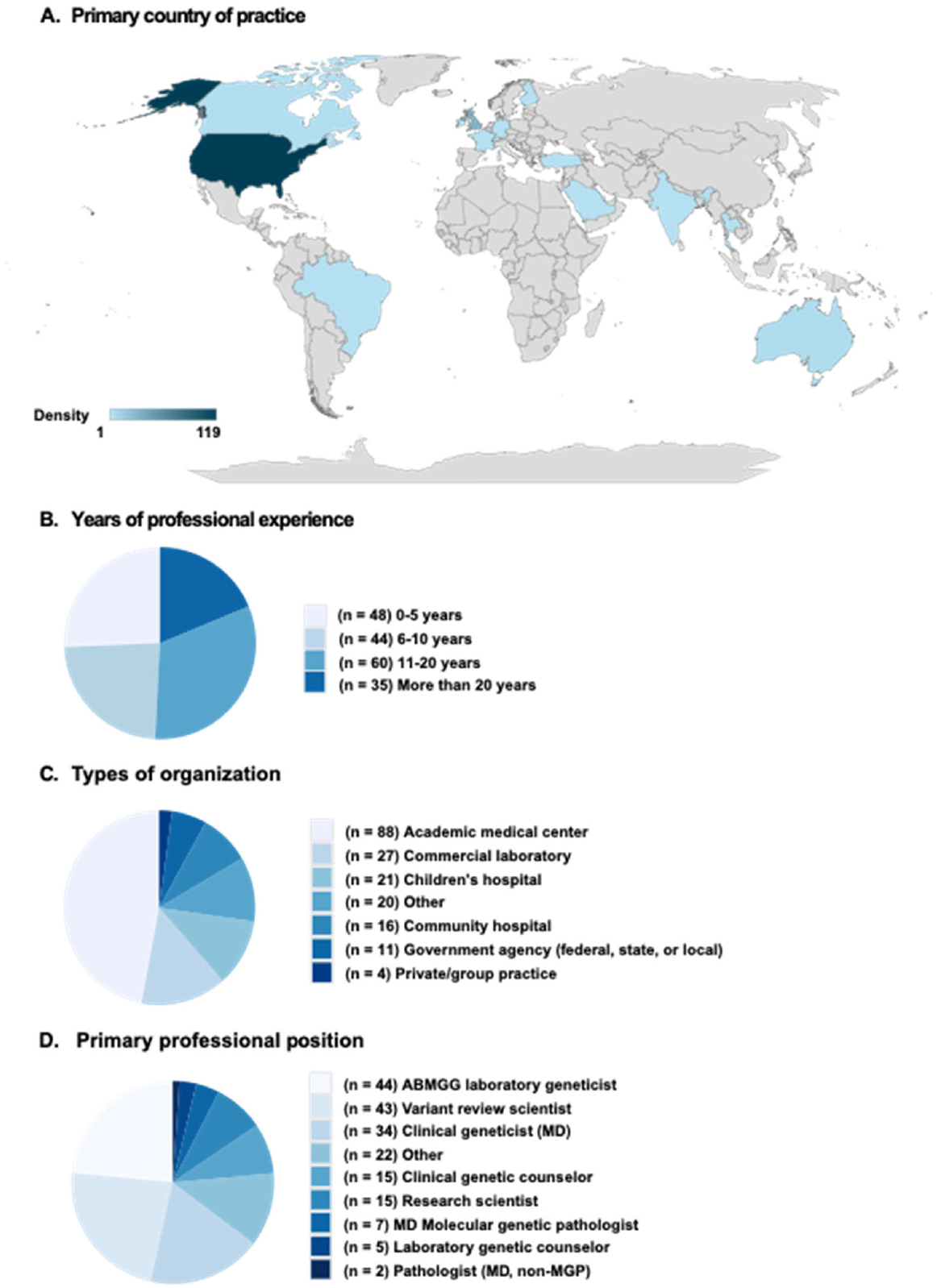
Respondents’ demographic information. **(A)** Respondents’ primary country of practice shown in a world map. The countries represented include United States, United Kingdom, Australia, Canada, Brazil, Finland, France, Germany, India, Ireland, Saudi Arabia, Singapore, Switzerland, Thailand, and Turkey. (n = 170); **(B)** Respondents’ years of professional experience divided into four groups. (n = 187); **(C)** Types of organizations respondents currently work for. (n = 187); **(D)** Primary professional role respondents hold. (n = 187)

**Figure 2.**
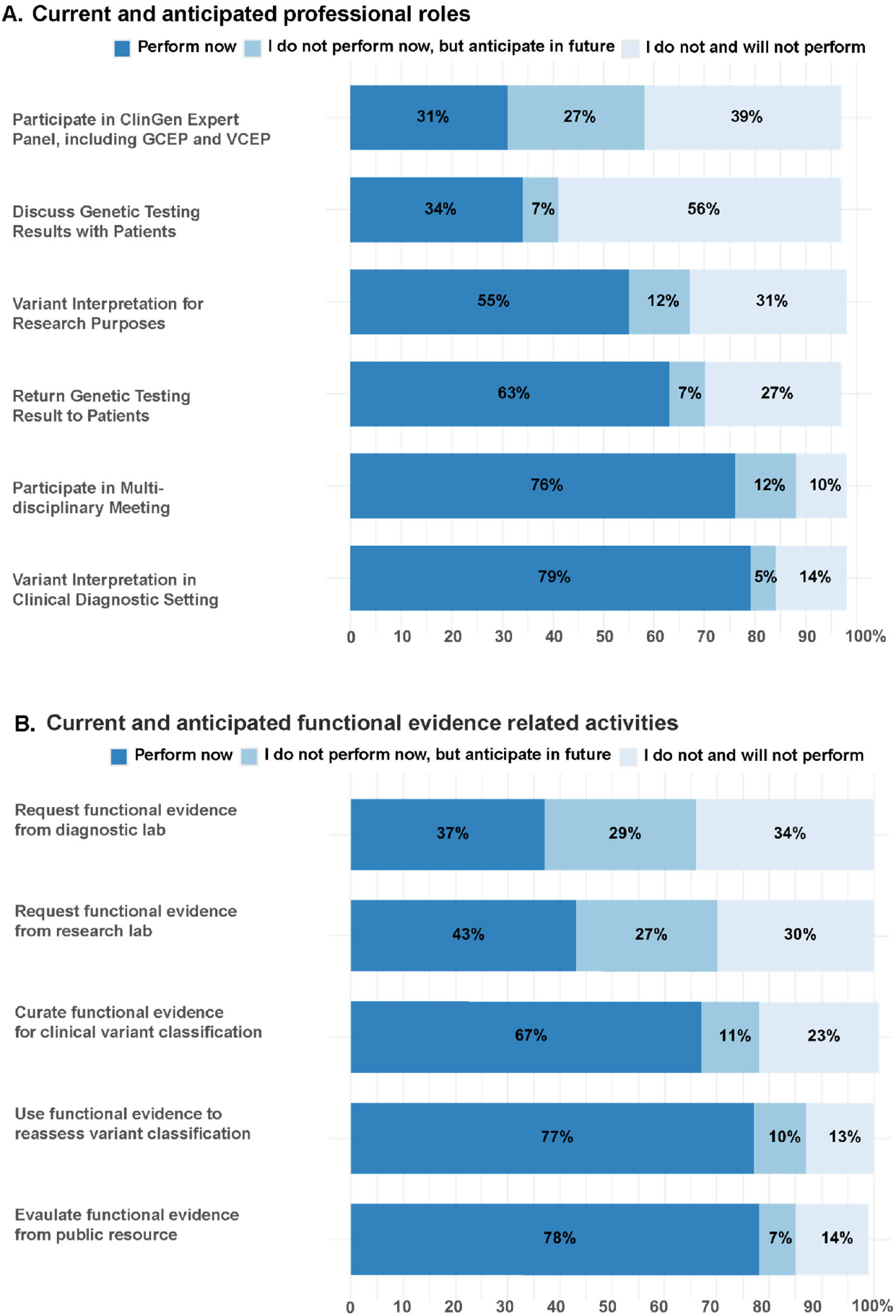
Proportion of respondents currently performing, planning to perform, or not performing various roles and activities. **(A)** Current and anticipated professional roles of respondents. (**B)** Current and anticipated participation in functional evidence related activities. N = 190 for both Figure 2A and 2B. Non-responses are not shown, but were less than 5% for all items.

### Insufficient evidence limits variant interpretation/reinterpretation

To understand the volume of VUS reinterpretations genome medicine providers faced, we asked how many VUS the respondents encountered in a typical year. Most reported interpreting 26 or more variants for clinical significance per year (92.6%) and more than a quarter reported *<u>re</u>*interpreting 26 or more VUS per year (26.8%) (**Figure S1A and S1B**). We then asked how often VUS were reinterpreted as B/LB or P/LP, and more than half reported that it occurs rarely or never (56.3%) (**Figure S1C)**. Not surprisingly, the majority of respondents reported that they interpret variants as VUS due to insufficient data frequently or very frequently (80.5%) and more than half reported that functional evidence data was rarely or never available for VUS (67.3%) (**Figure S1D and S1E**). Overall, this demonstrates that variant reinterpretation is a common practice in clinical genetics, but that reinterpretation is often limited by insufficient data.

### Comfort level with various functional evidence, guidelines, and resources for using functional data in variant interpretation

Functional data can be generated from a range of different assays, from traditional assays such as biochemical or splicing assays, to relatively new high-throughput assays such as MAVEs. We wanted to assess how comfortable respondents were when assessing results of various assays. While 90% of respondents reported being at least somewhat comfortable with biochemical assays, only 64% felt the same of high-throughput functional assays (**Figure 3**). Overall, respondents with more than 20 years of experience were more comfortable using functional evidence than respondents with 0-5 years of experience. However, irrespective of their years of experience a majority of respondents felt least comfortable with high-throughput assays out of the options listed, with 73% of respondents with more than 20 years of experience preferentially ranking high-throughput assays as their least comfortable functional assay compared to 59% of respondents with 0-5 years of experience (**Figure S2**).

**Figure 3.**
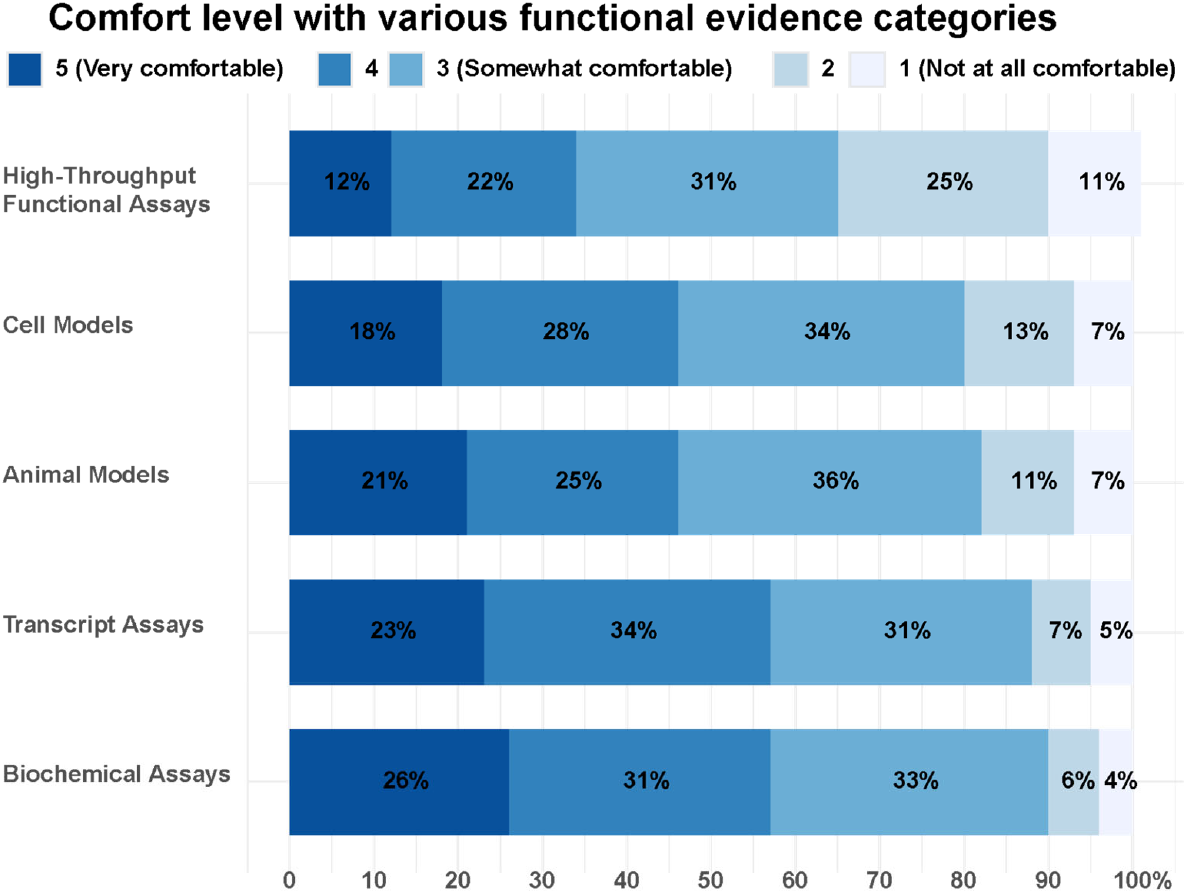
Respondents’ comfort level with various functional evidence categories. n = 190. Animal models had one missing value.

We also asked respondents about their knowledge of and comfort level with guidelines, databases and other resources available for using functional data. Among the functional evidence guideline options provided, the 2015 ACMG/AMP guidelines^4^ had the most respondents reporting as somewhat or more comfortable (91%) followed by 2020 updated recommendations for application of PS3/BS3 criterion for functional evidence (75%)^17^, whereas the ClinGen SVI’s functional assay assessment worksheet (https://clinicalgenome.org/docs/svi-functional-assay-documentation-worksheet/) had the lowest (23%; **Figure S3A**). Among the functional evidence resource options provided, primary literature had the most respondents reporting being at least somewhat comfortable with it (92%), followed by ClinVar and OMIM (69%; 65%). MaveDB, which is the database of record for high-throughput functional data^15^, had the fewest respondents reporting being somewhat confident with its use (20%), with a majority or respondents having not heard of this resource (51%; **Figure S3B**). Together, these findings demonstrate respondents are frequently unaware of, or not comfortable using existing resources outside of primary literature to aid incorporation of functional data into variant interpretation.

### Perceived barriers and facilitators of functional evidence use in variant interpretation

To better understand what factors can create barriers or facilitate use of functional evidence in variant interpretation, we asked respondents to consider a series of attributes for their helpfulness or unhelpfulness (**Figures 4A, 4B**). The two most frequently noted barriers were (1) insufficient information on the quality of functional evidence, and (2) insufficient confidence in the accuracy of the functional evidence, with more than half of the respondents reporting each of these to be barriers (61% and 55%, respectively). In the free text response section, almost half of the 19 responses mentioned challenge with interpreting the functional evidence for various reasons including uncertainty in appropriateness of the assay for a given mechanism of disease, validity of the assay, inconsistency in availability of functional evidence data, and time burden. One participant pointed out that “the evidence is NOT consistent, some genes, VUS have evidence, some has none, hard to decipher then how much emphasis to put on Functional Evidence in a busy Clinical practice”. Another response captures the time-consuming nature of assessing functional data for variant interpretation: “Time! Can only spend so much time on one variant/one patient.” Overall, these findings indicate that insufficient curation and validation of high-throughput functional assays is perceived as the major barrier for their use.

**Figure 4.**
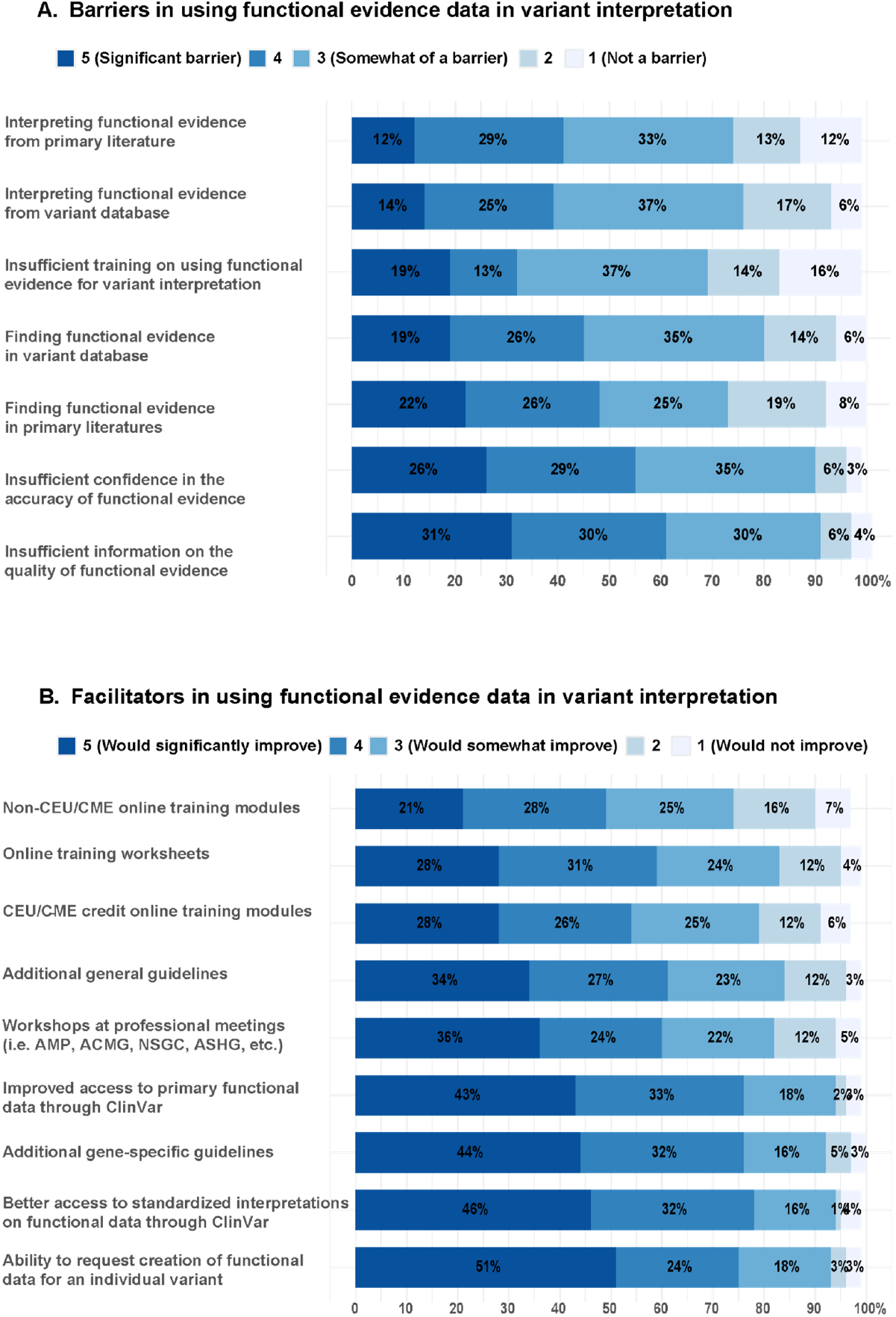
Respondents’ perception of potential barriers and facilitators for using functional evidence data in variant interpretation. **(A)** Degree to which (1 - not a barrier to 5 - significant barrier) respondents felt each of the options to be barriers to using functional data. **(B)** Degree to which (1 - would not improve to 5 - significantly improve) respondents felt each of the options to be facilitators for using functional data. n = 190 for both Figure 4A and 4B. Non-responses are not shown but were less than 5% for all items.

We also provided a list of resources and educational opportunities and asked respondents to assess their utility in improving respondents’ use of functional evidence data. Most respondents said improved access to standardized interpretation (PS3/BS3) of functional data (78%) and primary functional data through ClinVar (76%) would enhance their use of functional data (**Figure 4B**), highlighting the demand for better representation of functional evidence in ClinVar. Among the options provided, online training worksheets or modules either with or without CEU/CME credits were the least popular options for improving use (61%, 54%, and 49%, respectively; **Figure 4B**). These findings suggest that the respondents feel confident in their ability to assess functional evidence data if sufficient information and interpretation resources are provided.

### Attitudes towards a proposed functional data dashboard

In order to better understand how respondents would most prefer to interact with functional data, we presented a hypothetical dashboard for accessing MAVE-generated data, and asked them which features would be most helpful (**Figure 5**). Most respondents expressed overall enthusiasm for a centralized resource available through ClinVar. Specifically, over 84% of respondents agreed that inclusion of the following resources for variant-level functional data in ClinVar will improve their use of functional data in variant interpretation: (i) links to external websites to explore primary data;(ii) metrics for confidence/accuracy; and (iii) description of experimental design. One notable finding was a discrepancy in responses regarding including standard interpretation of the functional evidence (*i*.*e*., BS3/PS3 codes). As noted above, improved access to standard interpretations of functional data through ClinVar was one of the most popular options for promoting use of functional data (**Figure 4B**). However, including the standard interpretation was the least popular feature for a hypothetical dashboard, with 68% of respondents saying that they wanted this and 9% saying outright they would not use it for interpretations (**Figure 5**). Specifically, 16/190 (9%) respondents disagreed or strongly disagreed to using standardized interpretation (PS3/BS3) in their interpretation if included in the dashboard. However, 14 of those 16 respondents (88%) noted that access to standardized interpretation will improve their use of functional data. This indicates that although respondents want better access to curated interpretations of functional data, they are hesitant to use these curated interpretations without additionally having access to the functional data that support them.

**Figure 5.**
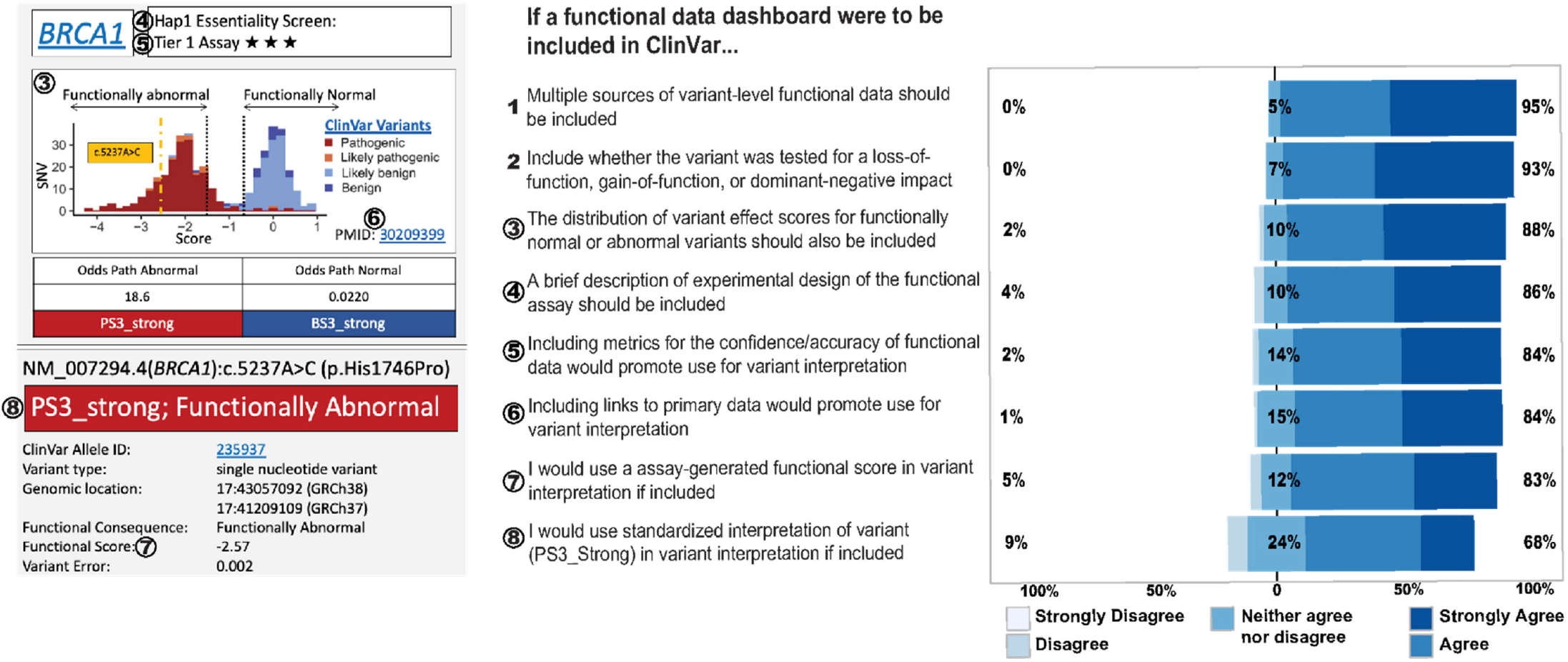
Respondents’ attitudes toward a potential functional data dashboard. Respondents were shown a hypothetical dashboard featuring MAVE-generated functional data for 4,000 *BRCA1* variants with various classifications (left). The dashboard included a histogram of MAVE scores, with black dotted lines indicating thresholds for functionally abnormal (left) and normal (right) variants. A yellow dotted line marked the score of an example variant. Below the histogram were ACMG evidence codes^17^, variant identifiers, the primary MAVE score, and associated measurement error. The respondents answered Likert-scale questions to assess their attitudes toward these features (right), circled numbers identify specific features of the dashboard.

### Handling conflicting functional evidence

The survey asked respondents if they had experience dealing with conflicting functional data, and allowed for free-text responses to elaborate on how. Three themes emerged from inductive analysis of the 116 responses (**Table 1**). 51% (59/116) of respondents described how they “assess the evidence.” While most conveyed the purpose of assessment was to identify the superior functional data for the disease of interest, the combination of factors considered in the assessment varied. Examples of factors mentioned in the responses include: consistency with patient presentation, presence of control data, internal validation, disease context, endorsement from expert committees, credibility of the primary literature, assay types, etc. Additionally, some respondents described that after the assessment, if they are still not able to determine which piece of data has more strength or weight, they either do not apply the PS3/BS3 criteria or take the conflicting data as a reason to classify the variant as VUS. One respondent’s answer captures concisely their decision-making process through the assessment and the complexity of it: “If both assays have clinical relevance, generally this can result in a non-application of functional weight. If one assay is superior in terms of clinical applicability and controls, sometimes override the conflicting data. Depends on the situation/assay”. The responses allude to the fact that assessing functional evidence for variant interpretation is subjective, complicated, and time consuming, with seven respondents specifically discussing the challenges of using primary literature for this process. The diversity of responses indicates that handling conflicting functional data is a part of variant interpretation that is non-systematically performed across institutions or labs.

**Table 1.**
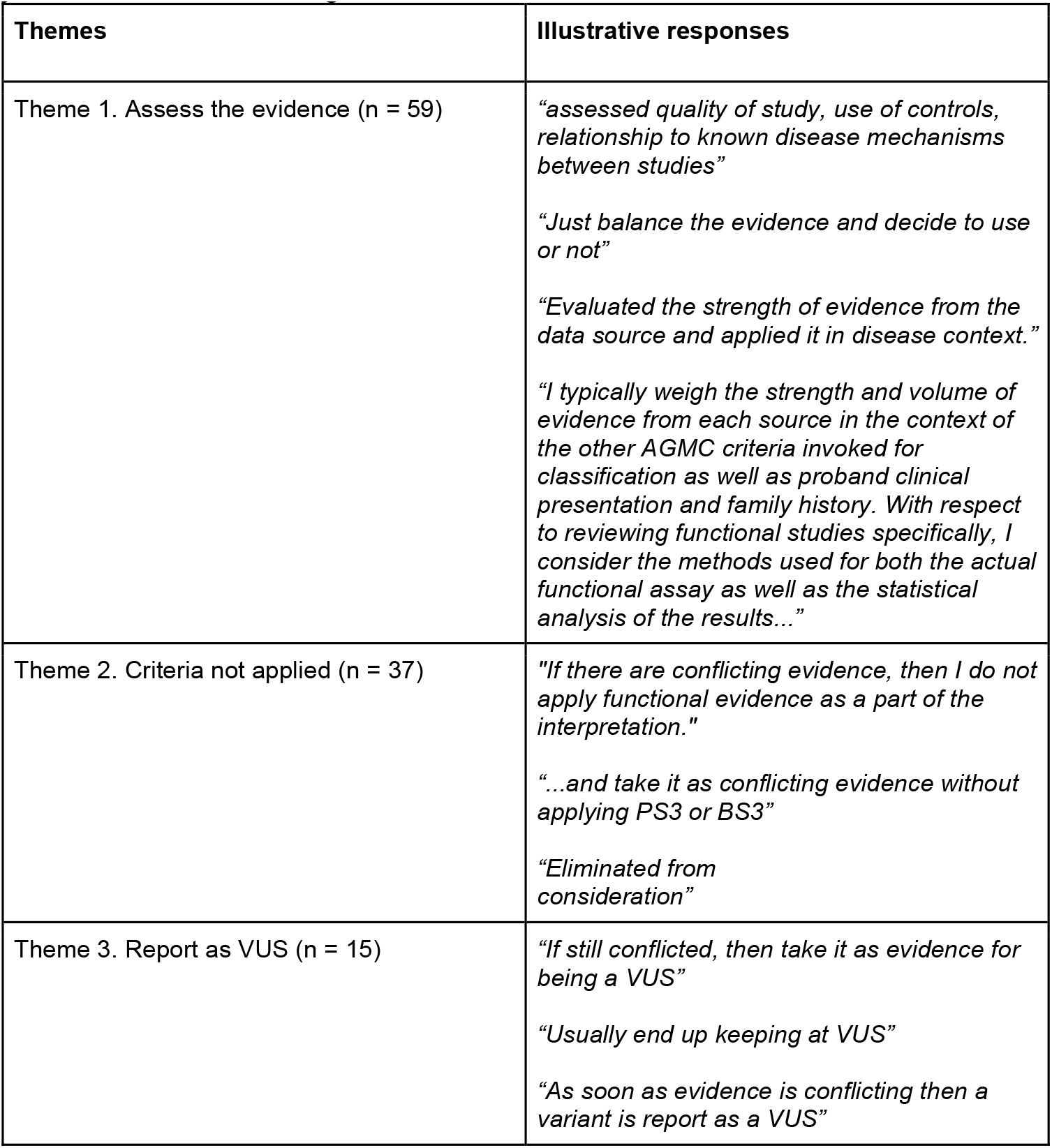
Qualitative analysis of 116 free text responses to the question *“Elaborate on how you have handled conflicting functional evidence”*.

| Themes | Illustrative responses |
| --- | --- |
| Theme 1. Assess the evidence (n = 59) | *“assessed quality of study, use of controls, relationship to known disease mechanisms between studies”*<br>*“Just balance the evidence and decide to use or not”*<br>*“Evaluated the strength of evidence from the data source and applied it in disease context.”*<br>*“I typically weigh the strength and volume of evidence from each source in the context of the other AGMC criteria invoked for classification as well as proband clinical presentation and family history. With respect to reviewing functional studies specifically, I consider the methods used for both the actual functional assay as well as the statistical analysis of the results...”* |
| Theme 2. Criteria not applied (n = 37) | *“If there are conflicting evidence, then I do not apply functional evidence as a part of the interpretation.”*<br>*“...and take it as conflicting evidence without applying PS3 or BS3”*<br>*“Eliminated from consideration”* |
| Theme 3. Report as VUS (n = 15) | *“If still conflicted, then take it as evidence for being a VUS”*<br>*“Usually end up keeping at VUS”*<br>*“As soon as evidence is conflicting then a variant is report as a VUS”* |

### Additional insights and comments provided by respondents

At the end of the survey, respondents were invited to share additional insights on functional evidence use in variant interpretation. Of the 20 who provided comments, the majority affirmed the importance of functional data in variant interpretation. Three respondents discussed incorporating functional evidence data into ClinVar: one preferred linking to a repository like MaveDB, while the other two supported making the data directly available on ClinVar. One respondent expressed the need for independently evaluating the evidence rather than relying solely on others’ interpretations and coming to their own conclusion since “[they are] ultimately responsible for the classification and interpretation in the clinical setting.” Another respondent mentioned that “additional clarity in […] how to apply functional [data] (more specifically MAVE data) to new ACMG guideline updates” will help determine the strength of the functional evidence data, which would provide support in applying the PS3/BS3 code.

In addition to expressing interest in improving guidelines, access, and availability of functional evidence, respondents also voiced frustration regarding the current status of affairs. One participant’s response effectively captured this sentiment: “I think I have to do entirely too much reclassification work […] Functional data, even when it exists, doesn’t always find its way into a report […] This is very frustrating for the clinician and families. Anything that can improve the mess we have now is welcome.”

## Discussion

The results from this survey provide insights into the current use of functional data in variant classification and factors that may further promote the use of functional data. Importantly, this survey captures a diverse set of providers who are actively involved in variant classification and reclassifications. In addition, 97% (184/190) of respondents completed at least 90% of the Likert-scale questions, and 61% (116/190) of respondents completed an optional free text question on how they handle conflicting functional data, reflecting respondents’ engagement with this 15-minute survey.

Respondents overwhelmingly reported that they currently use functional data in their practice but described gaps in their comfort level with different types of functional data. Specifically, respondents are universally the least comfortable with interpreting and using data from high-throughput assays for variant classification. As high-throughput functional data is on track to be the dominant functional data type used in variant interpretations, this survey exposes a significant barrier that needs to be addressed^25^. Importantly, this discomfort with high-throughput functional data applied to both early-career and late-career professionals, as well as both MDs and genetic counselors, indicating that this is a systematic gap across genetics providers.

Notably, insufficient training was not viewed as the most significant barrier to using functional data, and likewise, additional training was not viewed as the most useful method for improving the use of functional data for variant classification. Instead, respondents emphasized the need for improved access to standardized functional evidence data and clear guidelines for best practices for incorporating functional data into variant classifications, especially when presumably conflicting functional data exists. Specifically, respondents showed a strong interest in displaying functional data alongside variant-level data in databases like ClinVar. Furthermore, respondents emphasized the importance of including information that helped contextualize the functional data, including ranges of normal/abnormal values for the assay, whether conflicting data existed, experimental assay information, and disease mechanism information.

Surprisingly, although respondents overwhelmingly favored having standardized variant interpretations (*e*.*g*., indicating that PS3_strong has been met) in ClinVar and/or other repositories, these standardized interpretations on their own were viewed less favorably than that of the underlying data that support them. There may be multiple factors contributing to this, including the desire for provider autonomy^26^, as well as a lack of familiarity/confidence in current standards for converting functional data into evidence codes. However, irrespective of the underlying cause, this indicates that future work to incorporate standardized functional data into variant-level databases should prioritize inclusion of the underlying data to ensure provider uptake. This finding from our study, which predominantly included respondents practicing in the U.S., contrasts with a similar study conducted in Australia^27^. The Australian survey respondents appeared less confident in their ability to evaluate functional data. In contrast, respondents in our survey expressed less confidence in the underlying functional data itself, as well as the practices for converting functional data into variant classifications.

In addition, this survey exposed gaps in how providers handle conflicting functional data. Importantly, different assays may test different biological functions. Consequently, it is not necessarily biologically inconsistent to have two assays report out different effects for the same variant. For example, a functional assay that measures protein stability may report that a specific variant has “normal” protein stability, but a different assay that measures catalytic activity may report that a variant has “abnormal” catalytic activity. These functional effects are biologically consistent, but may appear as conflicting functional data. Teasing out which functional assay is the most appropriate to apply for variant interpretation - depending on the variant and the disease of interest - can be time-consuming and complex. In a busy clinical setting, this additional complexity may result in functional data not being utilized or the variant being interpreted or remaining as a VUS. Therefore, improving access to information on the appropriateness of specific assays for particular diseases could help promote the use of functional data, especially high-throughput assays, in variant interpretation. While only the qualitative aspect of this survey addressed how providers handle conflicting functional data, the breadth of responses to this open-ended question suggests that navigating such a situation may be a bottleneck in the use of functional evidence in clinical variant interpretation.

Overall, there appears to be a high degree of enthusiasm for a standardized and comprehensive resource for accessing functional data, but there are clearly barriers that need to be overcome to enable functional data to be systematically used in enabling precision medicine.

## Data Availability statement

The authors confirm that the data supporting the findings of this study are available within the article or at https://zenodo.org/records/14735954. Demographic data from the survey is available upon request, but is not on the zenodo above.

## Ethics Declaration

The study (#19511) was conducted in accordance with the declaration of Helsinki 1975, and in compliance with 45 CFR 56 for studies involving human participants. Waivers of consent, HIPAA, and involvement with minors were obtained from the University of Washington Biomedical Institutional Review Board (IRB).

## Conflict of Interest

The authors declare no significant conflict of interest.

## Acknowledgements

We thank the participants for taking the time to engage with this survey. We would also like to thank Rehan Villani and Mandy Spurdle for sharing their survey questions and Clare Turnbull and Sophie Allen for distributing the survey through CanVig-UK. This work was supported by an NIH NHGRI Advancing Medical Genomics Research award (R01HG013025) and LMS, DMF and AFR were supported by IGVF grant number UM1HG011969. ABS holds a Career Award for Medical Scientists from the Burroughs Wellcome Fund and is a Pew Biomedical Scholar. AEM was supported by an Early Career Award from the Alex’s Lemonade Stand for Childhood Cancer and *RUNX1* Foundation (21-25037) and a Brotman Baty Institute Catalytic Collaborations Grant (CC28).

## Author contributions

Analysis and Writing: MSP, RDK, LMS, ABS. Survey Design & Implementation: CO, AF, AM, AS, LMS, ABS. Study Conception: DMF, AFR, BHS, LMS, ABS

## Supplemental Figures

**Figure S1.**
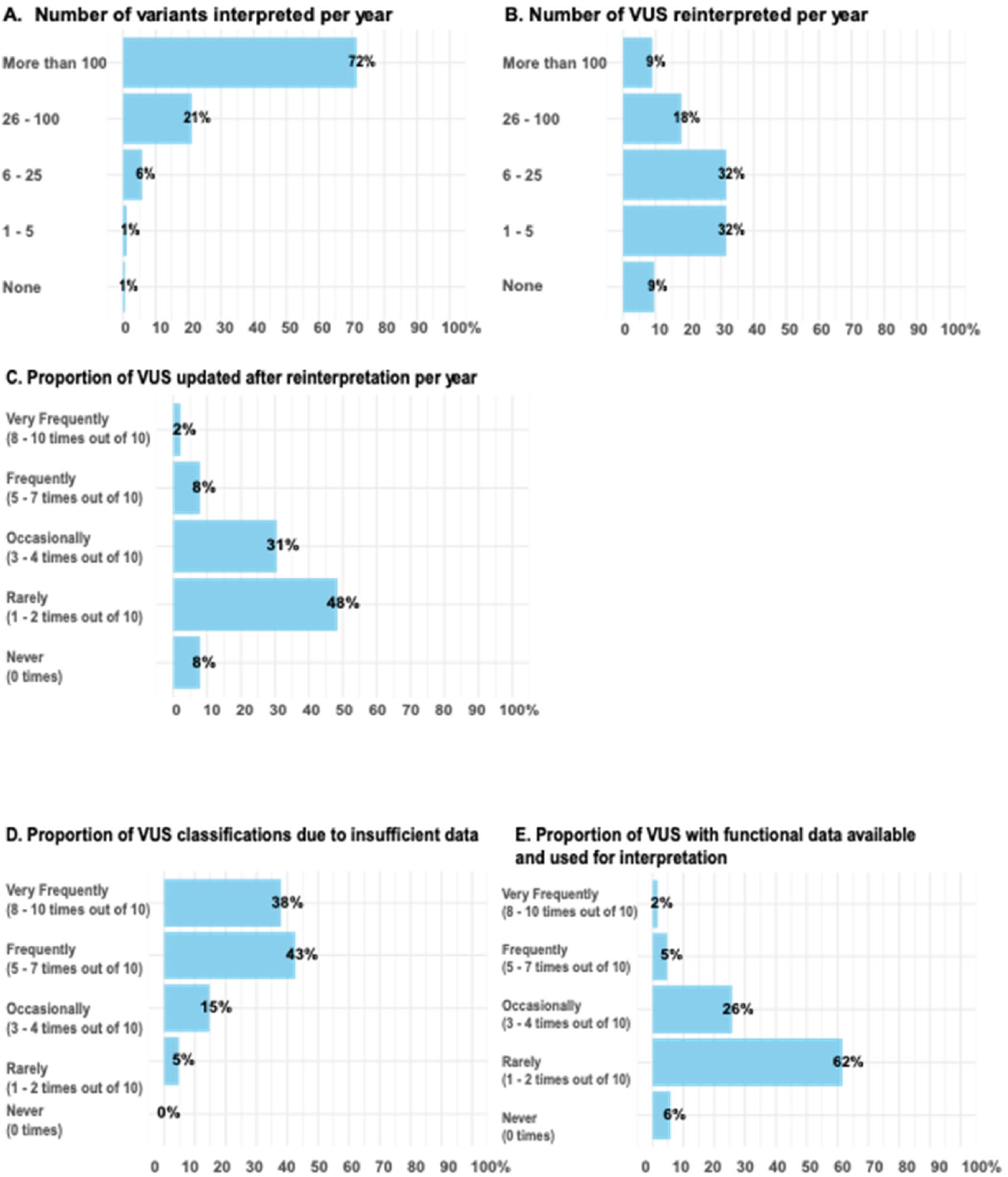
Exploration of variant interpretation practice by respondents. **(A)** Approximate number of variants interpreted by respondents per year divided into five groups (None, 1-5 variants, 6-25 variants, 26-100 variants, more than 100 variants). **(B)** Approximate number of VUS reinterpreted by respondents per year divided into five groups (None, 1-5 variants, 6-25 variants, 26-100 variants, more than 100 variants). **(C)** Proportion of VUS updated after reinterpretation by respondents per year. **(D)** Proportion of VUS classification that were due to insufficient data. **(F)** Proportion of VUS that had functional data available for use in variant interpretation. Scales ranging from “Never (0 times)” to “Very Frequently (8-10 times out of 10)” were used for Figure C, D, and E.

**Figure S2.**
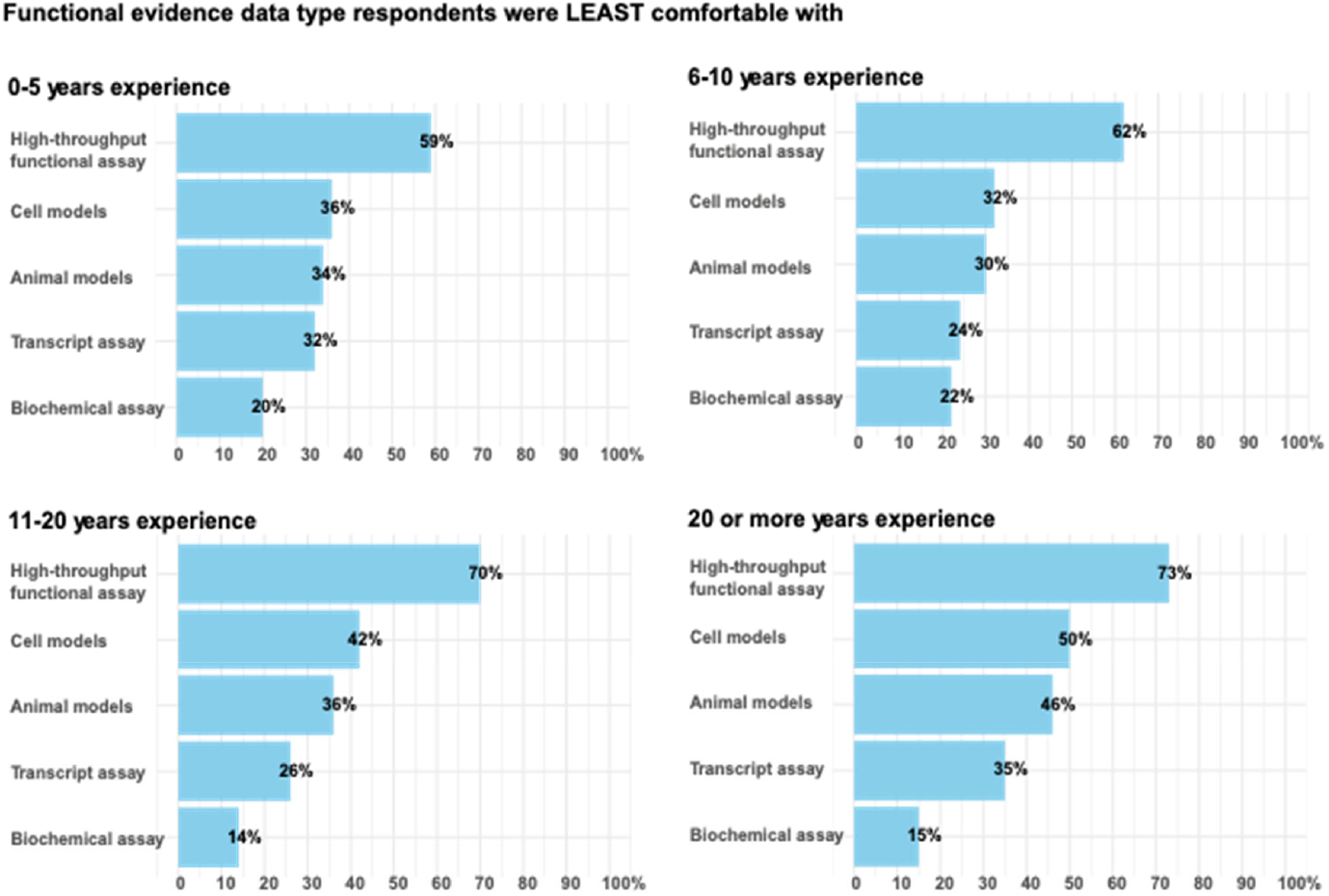
High-throughput functional assay as least comfortable functional evidence type across years of experience. Bar charts showing different types of functional assays ranked based on the comfort level score given by the respondents stratified by years of experience. Responses that gave the same score for all assays were excluded from this analysis. None of the responses were missing value for all five assays/models. Total n = 160 (0-5 years: n = 44, 6-10 years: n = 37, 11-20 years: n = 50, More than 20: n = 26).

**Figure S3.**
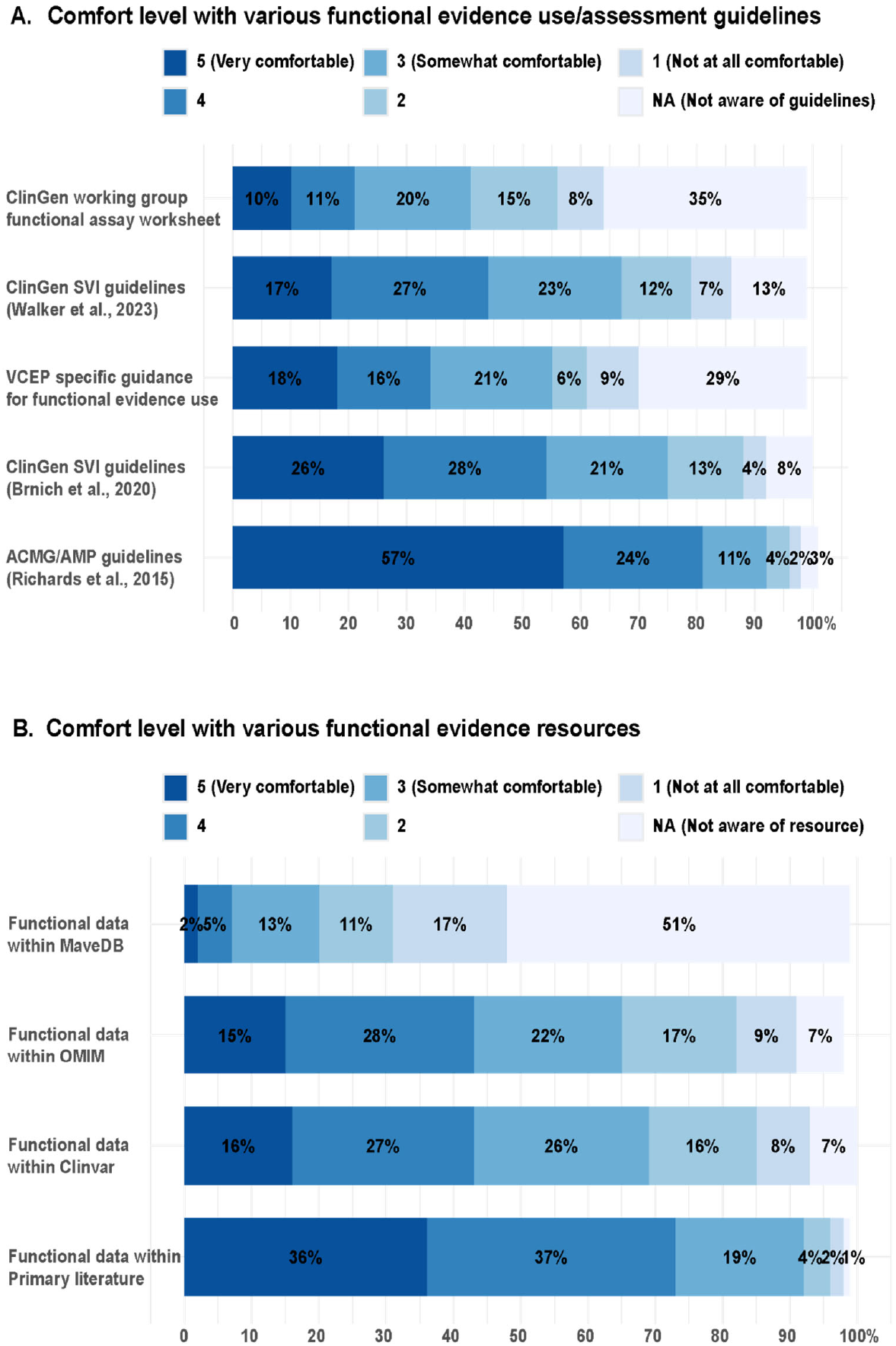
Respondents’ comfort level and awareness of various functional evidence use/assessment guidelines and resources. **(A)** Respondents’ comfort level and awareness of various functional evidence guidelines. (**B)** Respondents’ comfort level and awareness of various functional evidence resources. N = 190. Non-responses are not shown but were less than 2% for all items.

## Notes

### Competing Interest Statement

The authors have declared no competing interest.

